# Air sampling in team congregate spaces for early detection of respiratory virus threats at the 2026 FIFA World Cup™

**DOI:** 10.64898/2026.08.16.26360542

**Authors:** David Simon, Timothy J. Locksmith, Nicholas R. Minor, Isla E. Emmen, Nancy A. Wilson, Eli J. O’Connor, Shelby L. O’Connor, David H. O’Connor

## Abstract

**Background:** Respiratory infections are the leading cause of illness at major sporting events, yet surveillance relies on athletes recognising and reporting symptoms, missing asymptomatic, pre-symptomatic and unreported infections. During competitions, teams are housed together and share spaces with members of the broader community.

**Aim:** To evaluate whether continuous air sampling with point-of-care testing could detect respiratory-virus nucleic acids in an elite team’s congregate spaces during competition, and whether the signals were operationally useful.

**Methods:** Prospective, descriptive environmental-surveillance study following the Canadian men’s national soccer team across five host cities during the 2026 FIFA World Cup™ (3 June to 4 July 2026). InBio Apollo bioaerosol samplers ran continuously in up to four team-designated rooms per hotel (physiotherapy, meal, equipment and coaches’ room or hallway); filters were changed approximately twice daily, eluted on-site and tested with the Cepheid Xpert® Xpress SARS-CoV-2/Flu/RSV plus assay. A sample was considered positive if any cycle-threshold (Ct) value was reported, as less than 45, for a target.

**Results:** Of 174 air filters, 13 yielded detections of viral genetic material (9 SARS-CoV-2, 3 influenza A virus, 1 influenza B virus, 0 RSV). Detections were sparse early and clustered late in the tournament. An influenza A signal appeared the morning a player was sent home febrile, and SARS-CoV-2 signals coincided with visibly ill hotel staff, with signals falling after ill staff were excluded.

**Conclusion:** Air sampling with point-of-care testing is feasible in the context of an elite team and can surface behavior-independent viral signals during competition that may offer opportunities for earlier precautionary actions.

**Key public health message:** *What did you want to address in this study and why?:* Respiratory infections are the most common illness at major sporting events, but they are usually found only when someone notices symptoms and reports them. That misses infections in people who feel well or who choose not to say anything. We hypothesized that sampling and quickly testing the air of the shared rooms a national team occupies in a hotel would identify respiratory viruses, and that this could provide a behaviour-independent tool for improving elite athlete health.

*What have we learnt from this study?:* Air sampling with same-day testing worked in practice across five host cities and a month of competition. Most samples were negative, but viral genetic material was found in team rooms in every city. Some signals coincided with a febrile player and visibly unwell hotel staff.

*What are the implications of your findings for public health?:* Air sampling is a practical way to watch a shared indoor space for respiratory viruses without testing individuals or collecting personal data. When viruses are detected in a room, low-cost precautions such as ventilation, air cleaning or excluding unwell staff can be implemented. The same approach could be used for teams, delegations, and other groups housed together at mass gatherings in Europe and elsewhere.

## Introduction

Acute respiratory illness is the most common non-injury medical problem in elite sport and is consistently the single largest illness category at major sporting events. At the FIFA World Cup Qatar 2022™ it accounted for 80% (12 of 15) of all time-loss illnesses among players [1], and it was the leading illness category at the 2018 and 2020 Youth Olympic Games [2,3]. Elite athletes also carry elevated risk: at the 2019 Nordic World Ski Championships they had roughly a sevenfold higher risk of symptomatic respiratory infection than matched controls outside of the team [4], and infections spread readily within teams, most often within the same sport discipline [5,6].

Shared housing, communal meals, travel, and crowded indoor spaces are repeatedly identified as risk factors [7,8]. Team hotels concentrate these hazards, gathering susceptible athletes and staff in a closed, shared-ventilation environment. There, athletes and staff can be exposed both by one another and by other hotel occupants. Even if the athletes do not stray from the team environments, other hotel staff could bring viruses from the community into the team’s setting. The main infection route is inhalation of virus-laden aerosols, which accumulate and travel beyond close range in poorly ventilated air. Larger droplets can transmit virus during close face-to-face contact, and, less often, so can contact with contaminated surfaces [9].

Symptom-triggered surveillance is behavior-dependent, because infections are only ascertained when a person feels unwell, chooses to seek care or testing, and is correctly diagnosed. It misses or underestimates three situations that matter in team settings. First, asymptomatic and subclinical infections are common, with roughly a fifth of influenza and SARS-CoV-2 infections estimated to be asymptomatic [10–12], and RSV frequently goes unrecognised in adults [13]. Such infections are invisible to symptom-based monitoring yet can still shed virus and seed transmission.

The second class are symptomatic infections that are never reported or are reported after onset of symptoms. In one community study, only 17% (95% CI 10%–26%) of PCR-confirmed influenza infections received medical attention [14]. Even with highly conditioned athletes attuned to their bodies and close monitoring from trainers, during one professional rugby tournament, symptoms were present for at least a day before being reported to the team physician in more than half of illnesses [15]. In some cases, the incentive to keep competing may work against disclosure of symptoms.

Third, for many respiratory viruses shedding, and probably transmission, begins before symptoms do. This pre-symptomatic window is short, on the order of a day or two, but it is when an infected person is already seeding a shared space while still appearing well, which is why symptom-based detection lags the true course of a potential outbreak. In experimental influenza A infection most shedding preceded peak symptoms [16], influenza B shedding can rise up to two days before onset [17], and SARS-CoV-2 is detectable in the throat within roughly 40 hours of inoculation, before symptoms begin [18].

Elite team athletes cannot live in a pathogen-free bubble. Team members need to travel, hotel staff provide security and meal support, and team members mingle with family members who likely spend time in the surrounding communities. These individuals provide an avenue for a pathogen to infiltrate the team population [19,20].

A behavior-independent environmental signal such as viral nucleic acid in air could reveal a virus in a shared space occupied by people with asymptomatic, pre-symptomatic, or unreported infections. Respiratory viruses have long been recoverable from indoor air, and high-flow bioaerosol sampling is now a practical surveillance tool [21]. In congregate living settings, air samplers in student-dormitory HVAC returns detected SARS-CoV-2 in most cases when a positive resident lived on the same floor [22], and school air sampling has captured SARS-CoV-2 during sustained classroom transmission [23, 24]. Sampling across a range of community settings in Belgium, including nurseries, schools, workplaces, and nursing homes, detected at least one of 29 respiratory pathogens in 85% of 341 air samples, and showed that detection rose with higher carbon dioxide concentrations and poorer natural ventilation [25]. In a Belgian daycare centre, weekly air sampling over a full year combined with untargeted metagenomics recovered respiratory, enteric, and skin-associated viruses in 40 of 42 samples, with seasonal viruses appearing at epidemiologically expected times and complete genomes reconstructed directly from air [26]. Other work has detected viral nucleic acids in congregate settings such as schools and healthcare facilities [27] and at international airports [28].

To our knowledge, continuous indoor air sampling for early identification of viral threats has not been evaluated in the context of congregate elite athlete team communities. In partnership with Team Canada, we performed twice-daily air sampling in four team-hotel spaces during the 2026 FIFA World Cup™ to ask three questions: (1) can respiratory-virus nucleic acids be detected from the air of large, variably ventilated shared rooms that teams occupy; (2) are SARS-CoV-2, influenza A, influenza B, and RSV detectable during a summer competition, when Northern-hemisphere respiratory viruses circulate at low levels [29,30]; and (3) can air-sampling signals provide team physicians with behavior-independent information that is timely and specific enough to inform precautions?

## Methods

Reporting follows the STROBE recommendations for observational studies where applicable; because this is a descriptive environmental-surveillance and feasibility study without individual participants, items pertaining to individual-level exposures and outcomes are not applicable.

### Study design and setting

We conducted a prospective, descriptive environmental-surveillance study of respiratory-virus nucleic acids in the air of team congregate spaces occupied by the Canadian men’s national soccer team during the 2026 FIFA World Cup™. Sampling began on June 3, 2026 (during the final week of pre-tournament training) and continued as the team moved between host cities, ending on July 4, 2026 when the team was eliminated. In Montreal, Toronto, and Vancouver, samplers were placed in up to four team-designated rooms (physiotherapy, meal, equipment, and coaches’ rooms) at locations approved by the team physician. At the final two venues, room access was more limited, with three rooms sampled in Los Angeles and, in Houston, the coaches’ room was replaced by a team-occupied hallway. Room type rather than a fixed physical space was the unit of sampling, because rooms differed between hotels and varied widely in size and ventilation.

### Ethics and consent

This study sampled ambient air in shared congregate spaces and did not collect specimens from, or identifiers of, any individual. The University of Wisconsin-Madison Institutional Review Board has determined that air sampling in congregate settings, where the specific occupants of the sampled space are not known, does not constitute human-subjects research. Sampling locations were used with the agreement of, and coordinated with, Team Canada medical staff.

### Air sampling

Air was collected using InBio Apollo ambient air samplers (InBio), a filtration-based active bioaerosol sampler with a manufacturer-stated average flow rate of 540 L/min (≈32.4 ms/h) through its allergen-capture filter [31]. Samplers were positioned on available surfaces within each room and run continuously. These samplers are relatively low cost, at roughly 650 US dollars per unit, which made it practical to deploy several in parallel across the team’s shared spaces. They are also quiet (i.e., under 40 decibels) in operation, allowing them to be positioned discreetly in occupied rooms without interfering with team activities.

Filters were changed approximately twice daily, giving two collection sessions of roughly 8-12 hours each: a daytime session (∼07:00–08:00 to ∼14:00–16:00) and an overnight session (∼14:00–16:00 to ∼07:00–08:00). When a room was inaccessible (for example, when the equipment room was locked or the coaches’ room was in use), a single filter was run for ∼24 hours. Each filter carried a unique barcode; the sampler location, and the times of filter installation and removal, were recorded for every session. Occasionally, an additional sampler was added to a single room and was used as a second confirmatory air sample for the space.

### Sample processing and elution

All processing was performed on-site in a hotel room. Each filter was removed from its 3D-printed cassette holder with forceps and transferred to a tube containing 600–800 µL of phosphate-buffered saline with 0.1% Tween-20 (PBST); 800 µL was used typically. The tube was held at room temperature for ∼20 minutes to elute captured material, with intermittent agitation. The eluate was then transferred directly into a Cepheid Xpert® cartridge for testing.

### Molecular testing

Eluates were tested on the Cepheid GeneXpert® platform using the Xpert® Xpress SARS-CoV-2/Flu/RSV plus assay, a cartridge-based multiplex real-time RT-PCR that detects SARS-CoV-2, influenza A, influenza B, and respiratory syncytial virus (RSV). Each cartridge was run according to the manufacturer’s instructions.

### Definition of a positive detection

We defined a sample as positive for a given virus if the GeneXpert reported any cycle-threshold (Ct) value for that target, regardless of the instrument’s qualitative call. Because the Xpert® Xpress assay applies validated Ct cut-offs to return a qualitative “positive/negative” result optimized for clinical diagnosis from patient specimens, targets with a high Ct near the assay boundary may be reported qualitatively as negative by the instrument. For this type of air surveillance, where the goal is the earliest possible detection of viral signal rather than a clinical diagnosis, we treated any reported amplification as evidence of viral nucleic acid in the sampled air. This lower threshold appears appropriate, though it necessarily trades specificity for sensitivity. Each Xpert® cartridge includes an internal Sample Processing Control that verifies adequate processing. Samples with a non-amplifying control or a probe error, or otherwise determined to be invalid, were excluded from the figures but retained in the study data table.

### Target-enriched Illumina VSP2 viral sequencing

The final air-sample eluates from July 4, 2026 testing in Houston were used for targeted metagenomic sequencing. Nucleic acid from these eluates underwent target enrichment for over 200 respiratory viruses using the Illumina Viral Surveillance Panel v2 kit. Briefly, the samples underwent cDNA synthesis followed by a barcoding step with Illumina indices. The samples were batched together into groups of three, and then underwent a target hybridization reaction using an incubation time of 90 minutes. The capture was performed according to the protocol and a final amplification was done. Samples were then pooled together and sequenced on an Illumina NovaSeq. This sequencing was done by the UW-Madison Biotechnology Center.

Illumina paired-end reads were imported into Lungfish Genome Explorer (v0.5.0-beta25) using the VSP2 target-enrichment preprocessing recipe. On import, read pairs were adapter- and quality-trimmed and deduplicated with fastp (v1.3.2; sliding-window trimming, Q15, duplicate removal), depleted of human reads with deacon (v0.15.0) against the panhuman-1 index, and overlapping pairs were merged with fastp; merged and unmerged reads were then length-filtered to ≥50 bp with seqkit (v2.13.0) and deduplicated/reordered with BBTools clumpify (v39.80). The resulting single-end reads were analyzed with EsViritu (v3.14) against its curated viral reference database (v3.2.4), which applies its own quality filtering and read deduplication during competitive alignment to detect viral taxa and generate reference-guided consensus sequences. Because reads are deduplicated both at import (fastp/clumpify) and again during EsViritu mapping, reported read counts reflect unique reads per mapped taxa rather than raw sequenced reads.

### Outcomes and analysis

The primary output was the presence or absence of each of the four target viruses in each air sample. Ct values were used as a rough proxy for the amount of viral nucleic acid, with lower Ct indicating more material. No sample-size calculation was performed.

### Statistical analysis

Analyses were primarily descriptive. Detections were tabulated by virus, room, city, and sampling interval.

## Results

### Sampling overview

Over 32 days (June 3 to July 4, 2026), we collected and tested 179 air-sample filters for SARS-CoV-2, influenza A (IAV), influenza B (IBV), and RSV across five host cities: Montreal (n = 20), Toronto (n = 36), Vancouver (n = 79), Los Angeles (n = 9), and Houston (n = 35). Five filters (3 in Vancouver and 2 in Houston) were excluded from analysis because of invalid Cepheid GeneXpert tests. The remaining 174 filters were used for all analyses. Sampling covered four team spaces per hotel in Montreal, Toronto, and Vancouver (physiotherapy/treatment room, meal room, equipment/kit room, coaches’ meeting room), three in Los Angeles (no coaches’ room), and four in Houston (the coaches’ room was replaced with a team-occupied hallway). Sampling was briefly interrupted when teams and staffing equipment moved between each city.

Most samples were negative for all four targets. Across the study, 13 filters yielded a detection of at least one respiratory virus: SARS-CoV-2 in 9, IAV in 3, and IBV in 1. No RSV was detected. Detections and their cycle-threshold (Ct) values are summarized in Figure 1.

**Figure 1.**
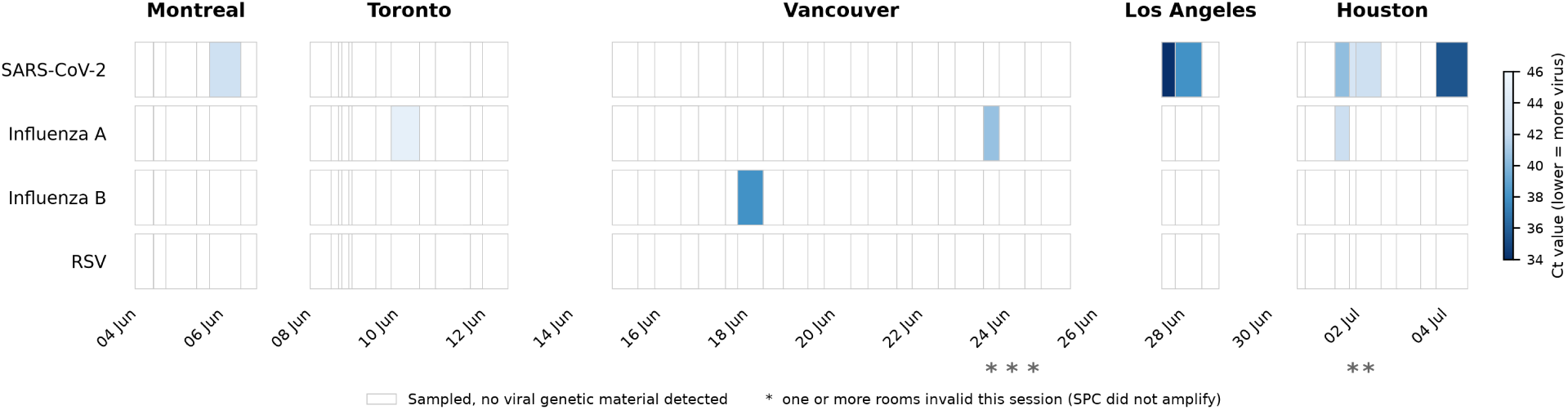
Respiratory-virus detections in team-space air samples across the 2026 FIFA World Cup™. Air was sampled continuously in Team Canada congregate spaces across five host cities from 3 June to 4 July 2026, and every filter was tested for SARS-CoV-2, influenza A, influenza B, and RSV. All cities are shown on a single continuous local-time axis. Rows are the four target viruses; each rectangle is up to 4 filters tested for one virus, drawn to its exact sampling window. The results from all rooms are merged within each city into a single rectangle. A box is blue if the virus was detected in any room (fill mapped to the lowest Ct across rooms), white if no room detected it but at least one returned a valid negative, and grey if every room was invalid. An asterisk beneath the axis marks a session where at least one room’s run was invalid and data were not available from all sampled rooms.

### Detections by city

Detections were sparse and geographically dispersed through the first three cities. In Montreal, SARS-CoV-2 was detected in the meal room on June 5 (Ct 42.8), against an otherwise negative baseline. Toronto also yielded one detection, when influenza A was detected in the physiotherapy room from the overnight session beginning June 9 (Ct 44.9). This value fell beyond the assay’s qualitative cut-off and was reported by the instrument as negative, but it met our detection definition of any reported amplification. In Vancouver, influenza B was detected in the physiotherapy room on June 17 (Ct 38.0) and influenza A was detected in the meal room on the morning of June 23 (Ct 40.6).

Detections became more frequent in the two final host cities. In Los Angeles, SARS-CoV-2 was detected in the meal room beginning on the first sampling day. On June 27, a filter was positive from the meal room in the daytime sampling interval (Ct 34.0). An additional filter was collected overnight from the meal room and also tested positive for SARS-CoV-2 genetic material (Ct 38.0). There were no additional positives before the air sampling supplies were moved on June 29 to Houston, where Team Canada played their Round of 16 match. Houston accounted for 7 of the 13 detections (Figure 2). On July 1, viral genetic material was found in the air of all four sampled areas: the meal room (Ct 40.4), hallway (Ct 42.1), and the physiotherapy room (Ct 43.4) tested positive for SARS-CoV-2, with influenza A in the equipment room (Ct 42.5). On the morning of July 2, only the meal room tested positive for SARS-CoV-2 from the previous overnight collection (Ct 42.7). No viruses were detected again until the morning of July 4 when SARS-CoV-2 was detected on samples at lower Ct values. There was a positive meal-room sample (Ct 35.5) and a positive hallway collection (Ct 37.8). One speculative interpretation, which we advance cautiously and cannot confirm, is that the July 1 signal reflected an introduction from one or more people from outside the team, the removal of the source individual(s) transiently reduced airborne virus, and that the lower-Ct resurgence reflected onward transmission within the team.

**Figure 2.**
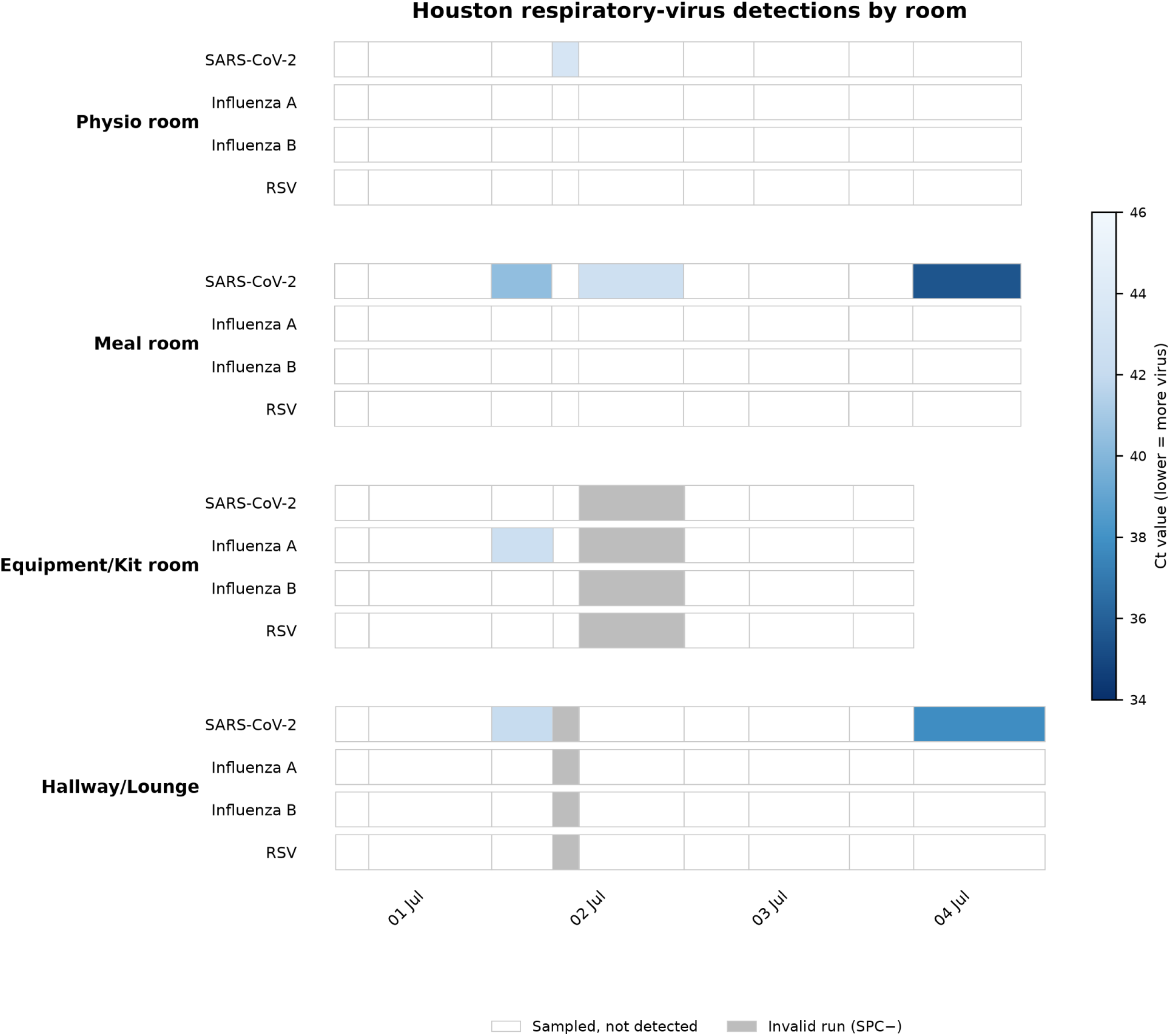
Per-room detections during the Houston sampling period. Sampling in Houston ran from the evening of 30 June to 4 July 2026 across four team spaces (physiotherapy room, meal room, equipment room, and a team-occupied hallway). Rows group the four target viruses within each room. White boxes are valid negatives, grey boxes are invalid runs, and blue boxes are detections (fill color mapped to Ct). On 1 July, SARS-CoV-2 was detected in the meal room, physiotherapy room, and hallway, and influenza A in the equipment room. SARS-CoV-2 returned on 3–4 July at lower Ct, most strongly in the meal room.

Community wastewater surveillance in all five host cities showed that the team’s visits fell in the seasonal trough for all four viruses, so the clustering of air detections in Los Angeles and Houston cannot be explained by unusually high background circulation (online supplemental methods and online supplementary figure 1) in these cities.

### Operational context and team responses

A prespecified aim was to determine whether air-sampling signals could give the team physician actionable, behavior-independent information. In Toronto, the influenza A detection in the physio-therapy room (overnight session from June 9, Ct 44.9) coincided with a player being withdrawn from training that morning with a fever. The team physician asked the player to mask and isolate for several days. In Los Angeles, after SARS-CoV-2 was detected in the meal room on June 27, medical staff asked visibly ill hotel personnel to leave the team spaces to reduce exposure, and no respiratory virus was detected in the subsequent June 28 samples, coinciding with the removal. In Houston, following the July 1 detections across multiple team spaces, medical staff again asked visibly ill hotel staff to leave, after which signal was absent on July 2 and returned on samples run from July 3 to July 4 at lower Ct values.

### Viral sequencing

The final four air samples that were being collected on July 4 (when Team Canada was eliminated from the tournament) were used for metagenomic sequencing with Illumina VSP2 target-capture, allowing us to detect additional human respiratory and environmental viruses across the sampled hotel spaces (Figure 3). In agreement with the Genexpert data showing SARS-CoV-2 genetic material in the Houston hotel between July 3 and July 4, SARS-CoV-2 sequences were detected from the meal room air sample collected on July 4, as well as from an equipment-room sample. Sequencing additionally recovered respiratory viruses not targeted by the Cepheid assay, human coronavirus OC43 and rhinovirus, alongside ubiquitous commensal viruses such as Merkel cell polyomavirus that are detected in nearly all sequences from human-occupied congregate spaces. No viral detections were obtained from the extraction blank or the no-template controls.

**Figure 3.**
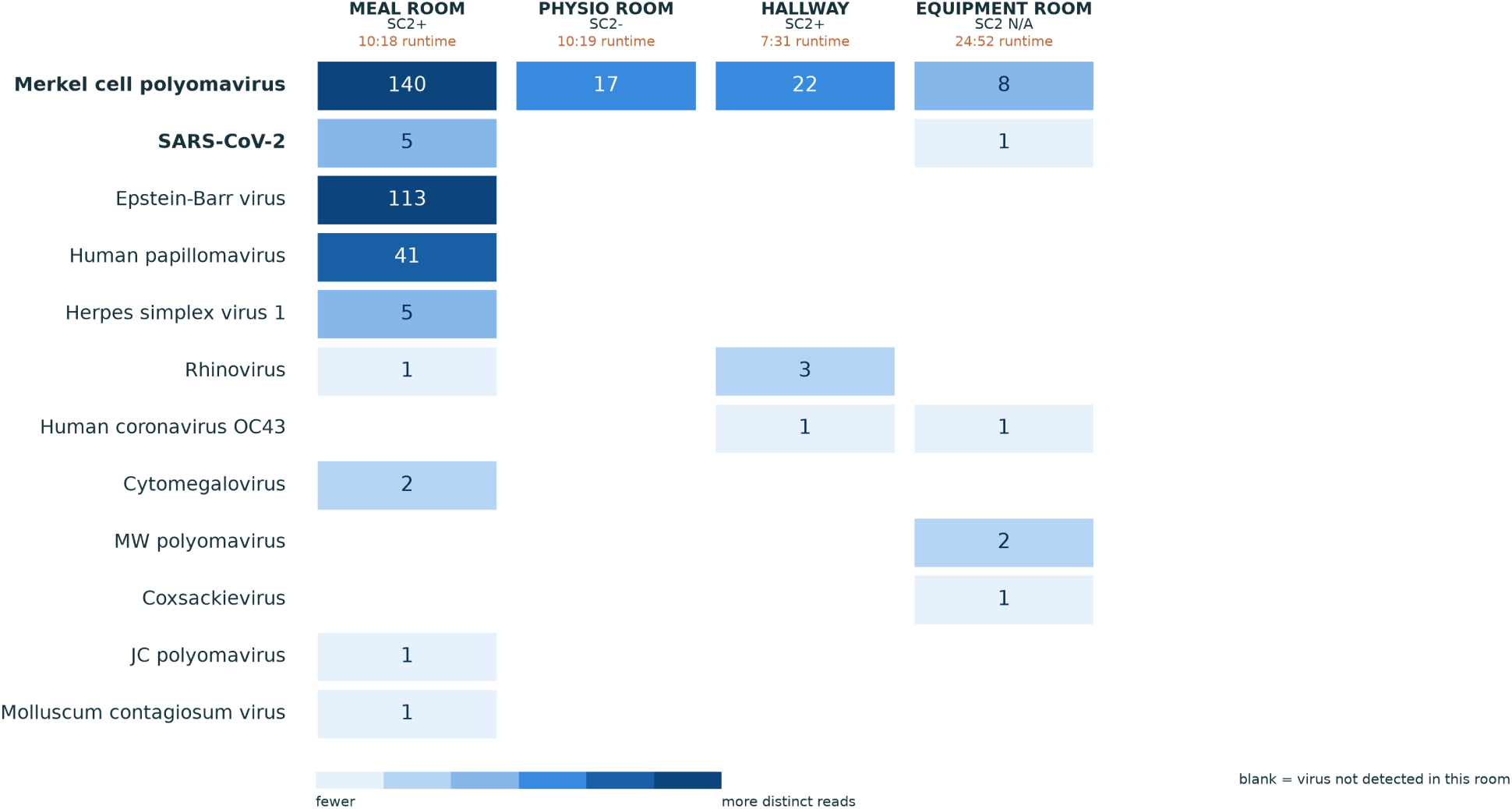
**Human viruses detected by metagenomic sequencing of the Houston air samples**. Air cartridges from the Houston team spaces were pulled from the samplers on the afternoon of 4 July 2026 and sequenced, and reads were classified against a curated viral reference database with EsViritu. These cartridges were collected the same day but are not the same cartridges tested on the GeneXpert, so sequencing provides an independent line of evidence for the viruses present in these spaces. Columns are the four sequenced samples, labelled by room, by GeneXpert SARS-CoV-2 status where a same-room GeneXpert result exists, and by the cartridge’s sampling runtime in hours and minutes (h:mm). Rows are the human viruses detected, with Merkel cell polyomavirus and SARS-CoV-2 placed at the top and the remaining viruses ordered by total distinct reads across all samples. Each cell reports the number of distinct (deduplicated) reads assigned to that virus in that sample, with fill mapped to that count on a light to deep blue scale, so deeper blue indicates more distinct reads. Merkel cell polyomavirus, a commensal virus of human skin, was found in every sample, consistent with continuous human occupancy. Sequencing additionally recovered respiratory viruses not on the GeneXpert panel, including human coronavirus OC43 and rhinovirus.

## Discussion

Figure 4 summarises the study design, the detections across all five host cities, and the response actions that a positive air signal can trigger.

**Figure 4.**
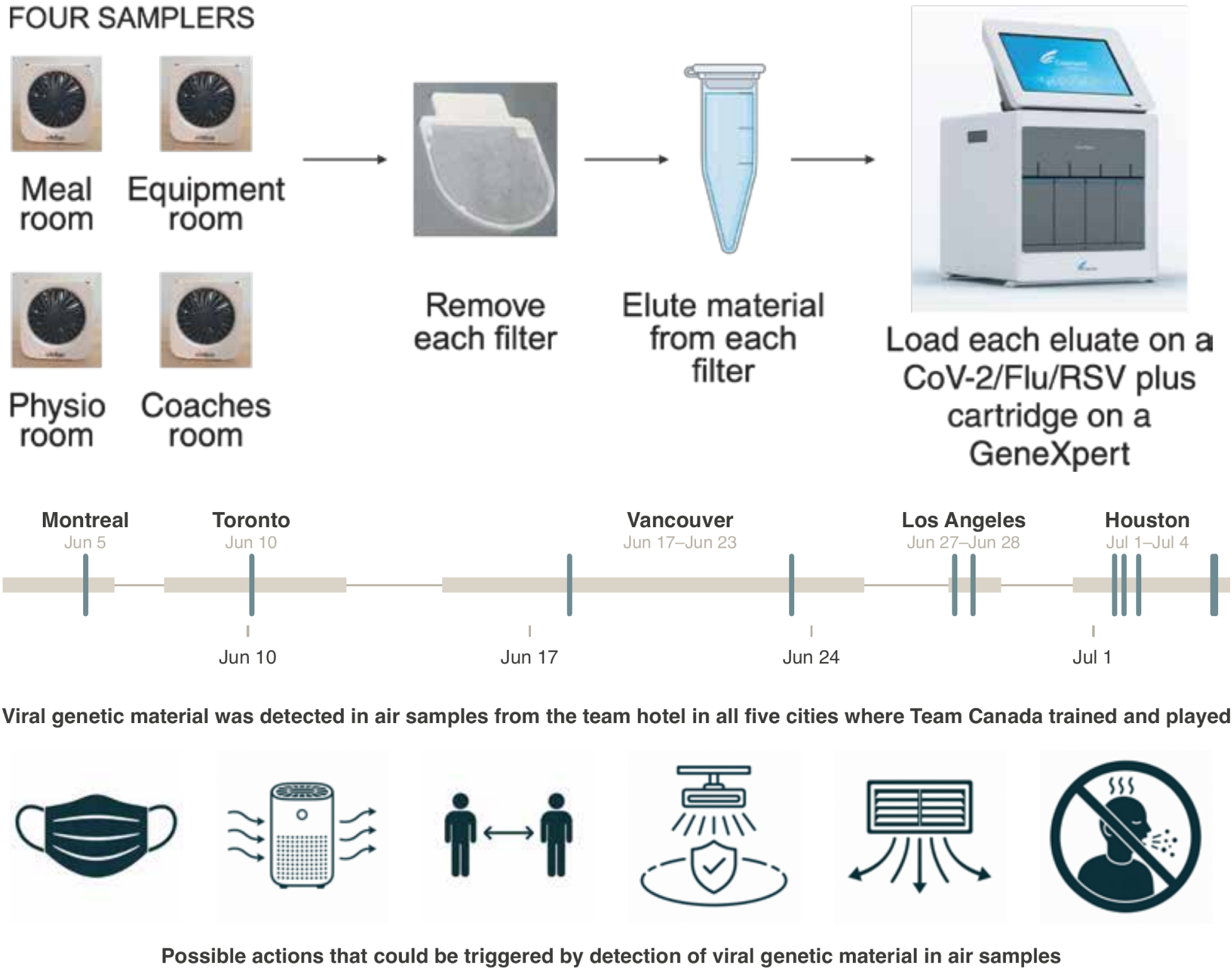
Air sampling for early detection of respiratory-virus threats in an elite team during competition. Continuous bioaerosol sampling was performed in up to four team congregate spaces per hotel, typically the meal, equipment, physiotherapy, and coaches’ rooms, across five host cities during the 2026 FIFA World Cup™. Each sampler’s filter was eluted on-site and tested on a Cepheid GeneXpert SARS-CoV-2/Flu/RSV plus cartridge, giving a same-day, point-of-care result (top). Viral genetic material was detected in team-hotel air in every one of the five cities. The timeline shows sampling coverage as a horizontal band and each viral detection as a vertical tick on a single continuous time axis across the tournament, with detections sparse early and clustered in the final cities (middle). A positive air signal can trigger low-cost, low-regret responses before an outbreak is clinically apparent, such as masking, portable air purifiers, distancing and reduced occupancy, far-UVC lighting, reinforced ventilation, and excluding visibly ill personnel (bottom).

Respiratory illness is consistently the single greatest illness burden in athlete health-surveillance programmes. Over four years of monitoring UK Olympic athletes it caused the largest share of 27,442 illness time-loss days [32], and it remains the leading medical problem at major Games even when prevention campaigns are in place, as at Paris 2024 [33]. Within a squad, congregate living amplifies risk. In a football academy, the SARS-CoV-2 attack rate was three times higher among those who both lived and trained on-site than among those who only trained there [34]. A single infection introduced into shared team spaces during a tournament can therefore threaten both athlete health and competitive availability.

Intensive infection-control measures unquestionably work in this population but are not sustainable indefinitely. Across four major winter-sport events, acute respiratory illness affected 38% of team members at PyeongChang 2018 and 26% at the 2019 World Ski Championships, but fell to 0% and 5% under the multilayered COVID-19 countermeasures at the 2021 and 2022 events, a roughly 10-fold reduction [35], and a parallel cohort showed annual illness incidence in elite skiers falling from 5.3 to 0.3 episodes per person during lockdown [7]. Those disruptive measures, such as quarantine, continuous masking, single-room housing, and restricted indoor facilities, are not compatible with normal competition across a month-long tournament, and illness returned as they relaxed [35]. An early-warning signal could instead let intensive precautions be applied selectively, escalated when risk is detected and relaxed when it is not.

For team medical practice, air sampling offers such a behavior-independent early-warning signal, one that does not depend on athletes reporting symptoms or submitting to individual testing. Even if team members are routinely tested, air sampling can assess whether other employees who occupy spaces in temporary hotel spaces could introduce viruses. This may be especially attractive in elite sports, where athletes may under-report illness to avoid being withheld from competition and where individual testing is logistically and ethically fraught during a tournament. A positive air signal can prompt low-cost, low-regret actions such as reinforcing ventilation [36], adding portable air purifiers [37], incorporating far-UVC lighting in highly trafficked spaces [38,39], masking in the implicated space, reviewing who has access to team areas, excluding visibly ill personnel, and heightening clinical vigilance.

The 2028 Men’s UEFA European Championship will bring 24 national squads to nine venues across eight cities in the United Kingdom and Ireland over a month of competition, and up to 12 of the 32 teams at the 2027 FIFA Women’s World Cup™ will qualify through UEFA. Each of those squads will occupy a succession of team hotels, sharing meal rooms, treatment rooms and equipment stores with staff drawn from each host community. Conducting similar air sampling within these teams could provide an innovative approach to reducing respiratory virus burden in highly consequential, limited duration competitions.

### Limitations

First, this is a descriptive, uncontrolled study of a single team. We observed associations between air signals and events on the team, but cannot establish that air sampling caused any reduction in transmission, nor that the operational responses were effective. Second, air detection demonstrates the presence of viral nucleic acid in a shared space. It does not identify who was infected, does not distinguish infectious virus from residual nucleic acid, and documents exposure rather than transmission. Third, our positivity definition (any reported Ct) increases sensitivity at the cost of specificity, and low-level signals near the limit of detection may include contamination or transient environmental nucleic acid. Fourth, the volume of air sampled per session, number of occupants in each space, and their distance from the air samplers are all uncertain, which limits any quantitative interpretation of Ct values. Finally, sampling was opportunistic and constrained by room access, competition logistics, and the team’s elimination, which ended data collection.

### Future research

The air samplers used here represent an early generation of the technology and are relatively insensitive, collecting onto a filter that must be removed and processed before any testing. The next generation of samplers is likely to integrate continuous, automated pathogen detection directly into the device. Programs such as the ARPA-H Building Resilient Environments for Air and Total Health initiative are explicitly developing autonomous indoor air biosensors that continuously monitor the biological content of the air and translate it into real-time risk estimates [40]. As these tools mature, approaches to early detection of respiratory threats in athletes should continue to improve in sensitivity, speed, and ease of deployment.

Assay breadth is likely to expand in parallel. Cepheid has described a GeneXpert Respiratory Panel prototype that runs on the same point-of-care platform used here and detects 26 respiratory pathogens, including rhinovirus, enterovirus, parainfluenza viruses, seasonal coronaviruses, adenoviruses, and metapneumoviruses alongside SARS-CoV-2, influenza, and RSV [41]. Applying a panel of this breadth to air samples would let air surveillance capture the common-cold viruses that account for much athlete illness but that our SARS-CoV-2, influenza, and RSV assay could not detect, giving better representation of overall respiratory illness burden.

## Conclusion

Twice-daily air sampling with same-day point-of-care testing was feasible in the mobile, high-pressure environment of a national team during a World Cup. Several detections coincided with a febrile player and with visibly ill staff, and prompted the team’s medical staff to act. These findings position congregate air surveillance as a promising, behavior-independent early-warning tool for protecting athlete health, and one that is directly transferable to the team hotels, delegations and congregate accommodation of mass-gathering events in Europe and elsewhere.

## Required end statements

### Ethical statement

The University of Wisconsin–Madison Institutional Review Board determined that air sampling in congregate settings, where the specific occupants of the sampled space are not known, does not constitute human-subjects research. The study did not collect specimens from, or identifiers of, any individual.

### Funding statement

This work was supported by Inkfish LLC and Heart of Racing.

## Data availability

The data and code for this study are available in the repository at https://github.com/dholab/team-canada-world-cup-2026. A compressed archive of the Lungfish Genome Explorer project (including imported sequencing reads) used to evaluate sequencing reads found in the July 4, 2026 samples can be accessed from https://dholk.primate.wisc.edu/_webdav/dho/public/manuscripts/team-canada-air-sampling/%40files/Team_Canada_VSP2.lungfish.zip. The original reads before import processing for Lungfish Genome Explorer are in NCBI SRA in Bioproject PRJNA1513008. Note that both the LGE and SRA datasets include data from two additional samples not discussed in the manuscript. During part of the tournament we tested an additional two air samplers for norovirus using the Cepheid GeneXpert and two of these samples that were running at the time of Team Canada’s elimination are included in the sequencing datasets. None of the air samples tested positive for norovirus on the GeneXpert; however, since we did not have complete coverage of the tournament we did not include it in the manuscript’s primary dataset.

## Confiict of interest

D.H.O. and S.L.O. are managing partners of Pathogenuity LLC, a consultancy that advises on topics including environmental monitoring for pathogens. D.H.O and S.L.O. are Honorary professorial fellows at the University of Melbourne, Australia.

## Authors’ contributions

DHO and SLO conceived and designed the study. DS facilitated access to team spaces, approved sampler placements, and provided clinical and operational context. SLO, EJO, DHO, TL, and NM conducted field air sampling, sample processing, and point-of-care testing across the host cities (SLO, EJO, and DHO in Montreal, Vancouver, Los Angeles, and Houston; TL in Toronto; NM in Vancouver). IE and NW performed the VSPv2 and uploaded the data to SRA. DHO and SLO drafted the manuscript, and all authors critically revised it for intellectual content, approved the final version, and agree to be accountable for all aspects of the work. Claude Opus 4.8 (high reasoning), Claude Opus 5 (high reasoning), and Codex Sol were used for background research, literature retrieval and review, manuscript text drafting and organization, data analysis and visualization, continuous integration of edits to produce the submitted versions of the text and figures, and adversarial simulated peer review. All LLM-generated content was independently verified and further edited by co-authors.

## Use of artificial intelligence tools

Claude Opus 4.8 (high reasoning), Claude Opus 5 (high reasoning), and Codex Sol were used for background research, literature retrieval and review, manuscript text drafting and organization, data analysis and visualization, continuous integration of edits to produce the submitted versions of the text and figures, and adversarial simulated peer review. All model-generated content was independently verified and further edited by the co-authors, who take full responsibility for the work.

## Data Availability

The data and code for this study are available in the repository at https://github.com/dholab/team-canada-world-cup-2026. A compressed archive of the Lungfish Genome Explorer project (including imported sequencing reads) used to evaluate sequencing reads found in the July 4, 2026 samples can be accessed from https://dholk.primate.wisc.edu/_webdav/dho/public/manuscripts/team-canada-air-sampling/%40files/Team_Canada_VSP2.lungfish.zip. The original reads before import processing for Lungfish Genome Explorer are in NCBI SRA in Bioproject PRJNA1513008.

https://dholk.primate.wisc.edu/_webdav/dho/public/manuscripts/team-canada-air-sampling/%40files/Team_Canada_VSP2.lungfish.zip

## Acknowledgements

We thank the players and staff of Team Canada for their cooperation and for accommodating air sampling in their team spaces during the 2026 FIFA World Cup™.

---

**Online supplemental figure 1.**
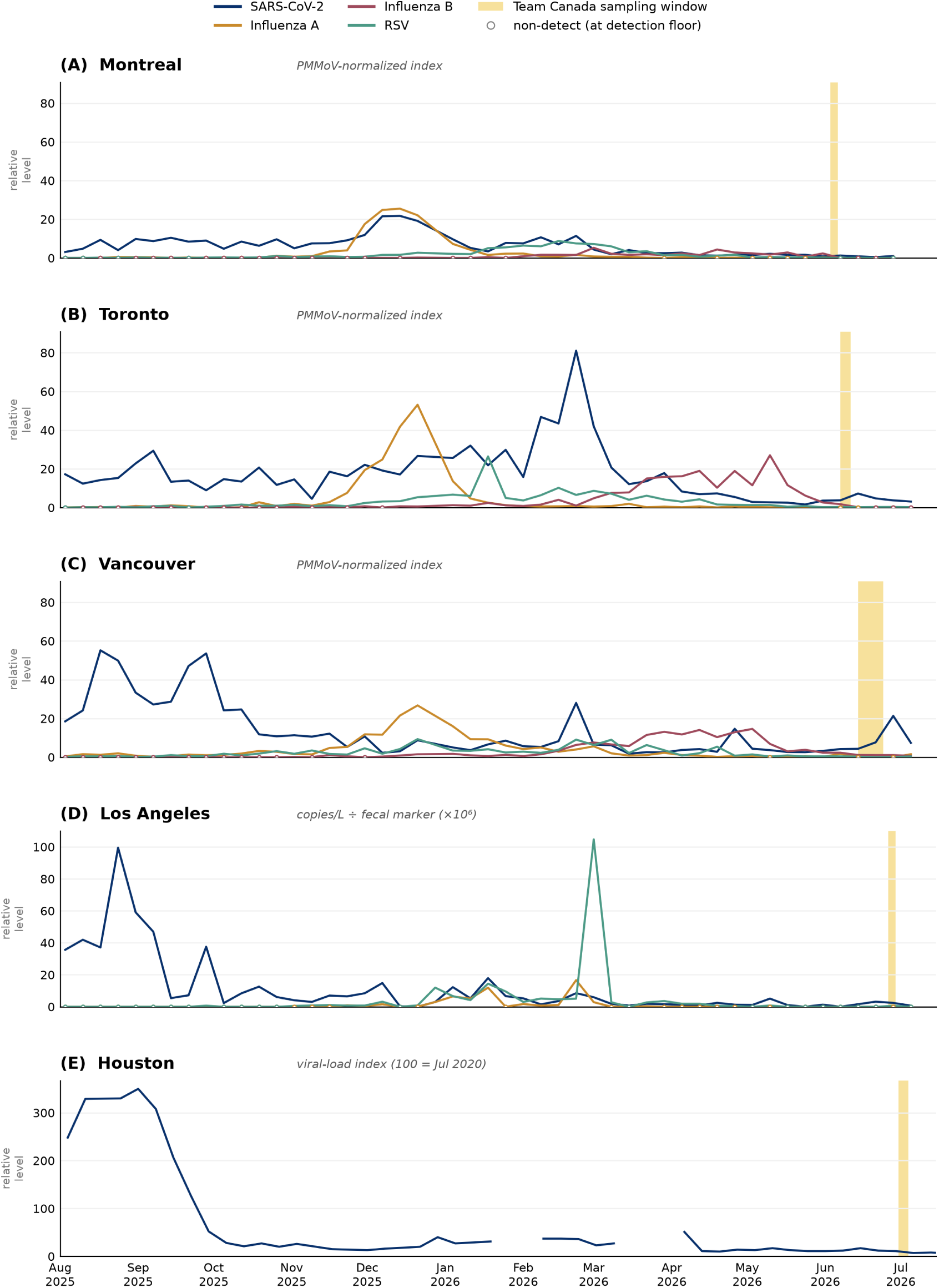
Community wastewater surveillance context in each host city over the 2025–2026 respiratory season. Each panel (A to E) is one host city, plotting publicly reported municipal wastewater levels for the four study targets (SARS-CoV-2, influenza A, influenza B, and RSV) from August 2025 through mid-July 2026. The shaded gold band marks Team Canada’s air-sampling window in that city. The four data sources report non-interchangeable quantities and are each shown on their own independent vertical axis; levels must be read within a panel and not compared between panels. Panels A–C (Montreal, Toronto, Vancouver) use the PHAC wastewater aggregate (PMMoV-normalized index); panel D (Los Angeles) uses the California CDPH/CDC-NWSS JWPCP dataset, self-normalized to fecal load; panel E (Houston) uses the Rice/Houston Health Department 69th Street index. Across all five cities the visit windows fall in the seasonal trough, at or near each site’s own lowest values, indicating no host city was experiencing unusually high community respiratory-virus circulation while Team Canada was present.

## Supplementary methods: community wastewater comparison

To place the air-sampling detections in the context of community-level respiratory-virus circulation in each host city, we retrieved publicly reported municipal wastewater surveillance data for the four study targets, covering the full 2025–2026 respiratory season (weeks beginning on or after August 1, 2025). For the three Canadian cities we used the Public Health Agency of Canada wastewater aggregate (Government of Canada Health Infobase, [42], accessed July 2026), which reports a PMMoV-normalized viral index for SARS-CoV-2 (measure covN2), influenza A, influenza B, and RSV. Vancouver is reported under the label “Metro Vancouver”, and where multiple sub-sites contributed we took the weekly median. For Los Angeles we used the California open wastewater dataset (California Surveillance of Wastewaters Network, California Department of Public Health and CDC National Wastewater Surveillance System, [43], data at [44], accessed July 2026) for the Joint Water Pollution Control Plant (JWPCP, Carson), the treatment plant serving the South Bay including Torrance, California, where the team was housed. Because that source reports absolute concentrations, we self-normalized each target to fecal load by dividing the target concentration (copies/L) by the concurrently reported human fecal marker concentration. Panel E (Houston) uses the Rice University and Houston Health Department dashboard for the 69th Street plant serving the downtown Main Street hotel, reporting the published viral-load index for SARS-CoV-2 only at plant level, scaled so that 100 represents the July 2020 baseline.

These four sources report non-interchangeable quantities on different scales, so the data cannot be compared between cities. Instead, each city is plotted on its own, with the team’s sampling window marked, and the comparison is strictly within-site, asking whether the levels the team sampled were unusually high relative to that same site’s own recent history. Non-detect values in the Canadian aggregate (reported at fixed detection-limit substitution values) were treated as non-detections. At plant level the Houston public feed reports SARS-CoV-2 only. The full per-city season is shown in online supplemental figure 1.

## Notes

### Summary of Updates

Updated to match a new submission to a different peer-reviewed journal. Minor text and stylistic edits.

